# Predictors and outcomes of Cardiac Dyssynchrony among patients with heart failure attending Benjamin Mkapa Hospital in Dodoma, central Tanzania: A protocol of prospective-longitudinal study

**DOI:** 10.1101/2023.06.15.23291476

**Authors:** Patrick Bilikundi, Baraka Alphonce, Azan Nyundo, John Meda

## Abstract

**Introduction:** Cardiac Dyssynchrony is prevalent among patients with heart failure with high cost of care and potentially poor outcomes. Nevertheless, little is known about cardiac dyssynchrony among heart failure patients, especially in developing countries. This study aims at assessing the predictors and outcomes of cardiac dyssynchrony among heart failure patients attending the cardiology department at Benjamin Mkapa Referral Hospital in Dodoma, central Tanzania

**Methods:** The study will follow a prospective longitudinal design involving participants aged 18yrs and above with heart failure attending the Cardiology Department at Benjamin Mkapa Hospital. Heart failure will be identified based on Framingham’s score and patients will be enrolled and followed up for six months. Baseline socio-demographic and clinical characteristics will be taken during enrollment. Outcomes of interest at six months include worsening of heart failure, readmission and death. Continuous data will be summarized as Mean (SD) or Median (IQR), and categorical data will be summarized using proportions and frequencies. Binary logistic regression will be used to determine predictors and outcomes of Cardiac Dyssynchrony among patients with heart failure.

## Introduction

Heart failure is a growing problem globally, resulting in increased mortality and morbidity[1,2]. Approximately 50% of individuals with heart failure die within 5 yrs of diagnosis[3], while yearly mortality is estimated at around 16%[1,3]. Others end up with recurrent hospitalization and worsening heart failure symptoms[4,5]. Cardiac dyssynchrony in the background of heart failure makes the outcome worse; up to 60% of individuals with heart failure and Cardiac Dyssynchrony die within four years of diagnosis[6].

Cardiac dyssynchrony is a phenomenon where there is delayed electrical activation in the cardiac myocytes (electrical dyssynchrony) or uncoordinated contraction of the cardiac ventricular walls (mechanical dyssynchrony), which can be either be between the segment of the cardiac ventricles (intraventricular dyssynchrony) or between the left and right ventricles (interventricular dyssynchrony)[7,8]

Prevalence of Electrical Dyssynchrony has been found to depend on the aetiology of heart failure [10]. Generally, it is as high as 11% in established heart failure patients, while in dilated cardiomyopathy is estimated to be 72.4%[8,9]. As for mechanical dyssynchrony, interventricular dyssynchrony has been found to be 79% while intraventricular is 75%[4,10]. Further studies have reported mechanical dyssynchrony in relation to QRS width, mechanical Dyssynchrony was found to be 20% interventricular and 40% interventricular, in those with QRS ≤ 120ms, while it was 58% intraventricular and 76% interventricular among those with QRS ≥120ms[11]. Moreover, Nagueh et al in Nigeria found that mechanical dyssynchrony in heart failure patients was as high as 27% for those with narrow QRS complex, while it was as high as 78% for those with wide QRS complex[9]. Additionally, among patients with heart failure; 34% prevalence of mechanical dyssynchrony was found in individuals with QRS ≥120 mms, and 11.9% in those with QRS of ≤120ms[12]. In another study that was conducted in Ivory Coast, among patients with dilated cardiomyopathy, mechanical Dyssynchrony was estimated to be 70% for intraventricular Dyssynchrony and 47.5% for interventricular Dyssynchrony[13].

Cardiac dyssynchrony in heart failure results in poor outcome, with increased in hospital readmission, worsening of heart failure and death [14]. Recurrent hospitalization and death were some of the outcomes reported, with prevalence of 37% and 19%, respectively[14] The development of these poor outcomes is multifactorial[5,13],advanced age, reduced ejection and NYHA class, especially class III and IV are among the key predictors of worse outcomes[15].Furthermore, left ventricle size (dilatation), heart failure etiology, especially dilated cardiomyopathy and ischemic heart disease, poor blood pressure control, diabetes mellitus and electrical dyssynchrony as defined by electrocardiogram are predictors for the development of mechanical dyssynchrony[16]

Studies done in Tanzania mainly focused on contemporary aetiology, clinical characteristics and outcomes of HF per se without considering Cardiac Dyssynchrony. For example, in a study which Makubi et al did was assessing the aetiological and common presenting symptoms of patients with heart failure, showed that hypertension was the main aetiology of HF, which accounted for 45% and 48% of the patients had New York Heart Association (NYHA) class III[17].

Despite few studies conducted in Nigeria and Ivory Coast on dyssynchrony in heart failure patients, little is still known about cardiac dyssynchrony in our setting, especially its morbidity and mortality, and as a result patients with heart failure and dyssynchrony miss the opportunity for appropriate intervention such as Cardiac resynchronization therapy (CRTDs). This will offer convincing justification for developing therapeutic services (CRTDs) in our area, incorporating them into local guidelines, and persuading the health insurance companies to include them in their packages.

Therefore, this study aims to determine the prevalence, predictors and outcome of cardiac Dyssynchrony among patients with heart failure who will be attending the Cardiology clinic at Benjamin Mkapa Hospital in Dodoma, central Tanzania

## Materials and Methods

### Study aims

1. To determine the prevalence of Cardiac Dyssynchrony among adult patients with heart failure attending Cardiology clinic at Benjamin Mkapa Hospital in Dodoma, Tanzania.
2. To determine the predictors of cardiac Dyssynchrony among patients with heart failure attending Benjamin Mkapa Hospital in Dodoma, Tanzania.
3. To determine the six-month outcomes of patients with heart failure and cardiac Dyssynchrony attending Benjamin Mkapa Hospital in Dodoma, Tanzania.

### Study design

A prospective longitudinal observational study will be conducted among adult patients with heart failure attending cardiac clinic at Benjamin Mkapa hospital.

### Study area

The study will be conducted in the cardiology department at the Benjamin Mkapa Hospital (BMH), located in the city of Dodoma, 16 kilometres from the city centre. BMH serves as a consultant hospital for the Central zone and other neighbouring areas of Dodoma. The hospital serves around 5.68 million people according to 2022 TDHS, majority being from Dodoma region, about 11.3million, while the rest are from nearby regions, including Singida, Manyara, Tabora, Morogoro, and Iringa[18].Moreover, it is used as a teaching hospital for the University of Dodoma (UDOM). This hospital helps in providing health care services for patients in the Central zone. Advanced radiological tests, including CT scans, MRIs, cardiological diagnostic tests, and interventional procedures like Coronary Angiography, Percutaneous Coronary Intervention, and Cardiac pacing are available at Benjamin Mkapa Hospital and are utilized to treat numerous patients from all over Tanzania.

BMH has a bed capacity of 400, and approximately 240 patients with heart conditions usually attend the cardiology department at BMH per month, of whom 15-20 patients are of Heart Failure and a six patients with Cardiac Dyssynchrony are registered per week from both BMH (Unpublished data).

### Sample size estimation

In order to estimate the sample size, the proportionate formula used in prospective cohort studies will be used[19]

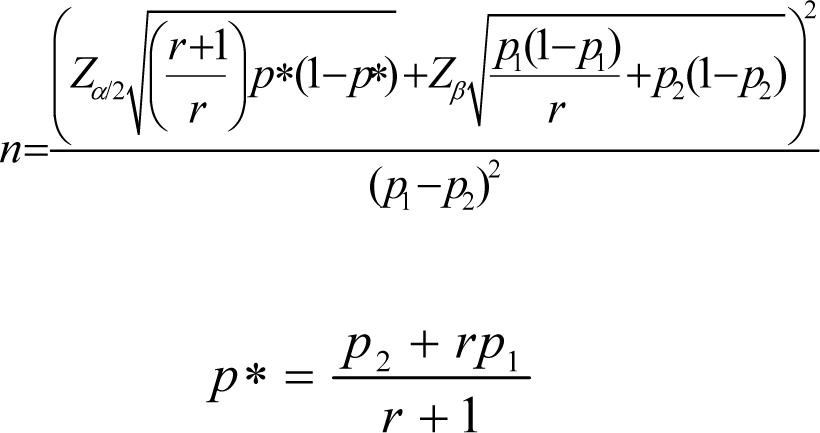

Where by

r = ratio between the two groups

p1 = Cardiac dyssynchrony prevalence obtained from literature p2 = expected prevalence from the study

p1 – p2 = effect size

𝑍_𝛽_ = standard normal variate for statistical power

𝑍_∝/2_= standard normal variate for significance level

The prevalence of cardiac dyssynchrony in previous studies was 12.9% The expected prevalence in this study is expected to be 16% Therefore;

r = 1

p1 = 12.9%

p2 = 16%

p1 – p2 = -3.1%

𝑍_𝛽_ = 0.84 for statistical power of 80%

𝑍_∝/2_ = 1.96 for significance level of 95% N= 134

Considering the 20 % attrition rate in our setting

Therefore, the minimum sample size estimated is 150 patients.

### Inclusion criteria

1. All patients aged ≥18 years with signs and symptoms of heart failure based on the Framingham score
2. All patients who are able to provide informed consent, and for those who will be unable a surrogate relative will consent on their behalf.

### Exclusion criteria

1. All patients with heart failure aged ≥18 years with LVAD.
2. All patients with heart failure aged ≥18 years with a functional pacemaker
3. All patients with heart failure aged ≥18yrs with ventricular rhythms.
4. All patients with advanced malignancies or end stage medical conditions

### Sampling methods/techniques

Consecutive Sampling method will be used whereby the sample will be attained by selecting every participant who fits the inclusion criteria until the desired sample size is reached.

### Assessment of heart failure

A detailed history and general physical examination will be conducted on all participants. The diagnosis of heart failure will be reached based on Framing Harm criteria; two major criteria, or one major and two minor criteria [20]. Major criteria will include: Paroxysmal nocturnal dyspnoea, Neck vein distention, Rales (crackles in the lungs) Cardiomegaly (enlarged heart) on chest X-ray Acute pulmonary oedema S3 gallop (a third heart sound). Minor criteria will include: Oedema, Nocturnal cough, Hepatomegaly (enlarged liver), Pleural effusion (fluid accumulation around the lungs) and Dyspnoea on exertion

### Assessment of Cardiac Dyssynchrony

#### Electrical Dyssynchrony

A standard 12–lead surface electrocardiogram (ECG) machine (GE Healthcare MAC^TM^ 2000 USA; 2020) set at a paper speed of 25 mm/s and a scale of 10 mm/mV will be used to assess electrical Dyssynchrony. The procedures for performing ECG will follow the standard operating procedure set by the cardiology department of the Benjamin Mkapa hospital. This protocol was adopted and customized from the established guidelines of the American Heart Association[21] Participant will be asked to expose the chest adequately and positioned supine on the cardiac bed. There will be no need to shave the skin as long as wet gel electrodes are used. Precordial leads will be six; V1-V6, V1 and V2 will be placed along the fourth intercostal space, 2cm right and left side of the sternum respectively. V4 will be placed in the fifth intercostal space, while V3 will be placed in between V2 and V4. V5 and V6 will be placed at fifth intercostal space, along the medial axillary line and axillary line respectively. The limb electrodes will be applied as follows: Red on the right inner wrist, yellow on the left inner wrist, black just above the ankle on the right inner leg, and green just above the ankle on the left inner leg. After entering the patient’s ID number into the device, a printout will be obtained. Electrical dyssynchrony will be ascribed to those with a widened RSS complex of more than 120ms[5]. Interpretation of the findings will be done by the principal investigator and confirmed by two experienced cardiologists. Any disagreement will be settled in agreement with a third independent experienced cardiologist.

#### Assessment of Mechanical Dyssynchrony

Mechanical Dyssynchrony will be assessed using Transthoracic echocardiography (model Vivid ^TM^ T9 made by GE Healthcare, USA 2018). The patient will be asked to expose the chest and abdomen above the symphysis pubis and positioned in the left lateral position. ECG tracing rhythm leads will be placed on the chest to pick the QRS complexes timing the onset of contraction corresponding to the aortic and pulmonic valvular closer. A two-dimensional (2D), M-mode, Doppler echocardiogram and speckle tracking echo will be performed; parasternal, short axis, apical four-chamber views and five chamber view. The dimensions of the left atrial, LV, and other cardiac chambers will be determined using a 2D guided M-mode approach. The left ventricle pre-ejection interval (LPEI), right ventricular pre-ejection interval (RPEI), and septal to posterior wall motion delay will be assessed[22].

The time between the start of the QRS complex and the start of the aortic flow velocity detected in the apical five-chamber image is used to compute the aortic pre-ejection time, termed as LPEI. From the start of the QRS complex until the start of the pulmonary flow velocity curve visible in the left parasternal view, the pulmonary pre-ejection time is calculated, termed as RPEI. LPEI will be defined as the distance measured from the start of aortic Doppler flow velocity curve to the start of QRS, while RPEI will be defined as the distance measured from the start of the pulmonary doppler flow velocity curves to the start of the QRS. A 40ms difference between LPEI and RPEI will be considered as inter ventricular mechanical delay (IVMD). The time interval between the maximal outward displacement of the inter ventricular septum and the LV posterior wall will be measured as a septal to posterior wall motion delay (SPWMD)[23] and a value of >130 ms 2 standard deviation above the mean of normal controls will be considered the presence of intraventricular dyssynchrony[9]. Intraventricular dyssynchrony will be defined as LPEI 140 ms in patients with difficult-to-identify systolic peaks of the LV posterior wall or an akinetic interventricular septum[24].

### Outcome measurement

#### Primary outcome

The primary outcome of interest will be the presence of cardiac dyssynchrony. Cardiac dyssynchrony will be either mechanical, electrical or both

##### Mechanical dyssynchrony

**An** echo will be performed to evaluate the delay in contractility in ventricular chambers between the chambers (interventricular) or between segments of the left ventricle (intraventricular)

##### Electrical dyssynchrony

a standard 12 ECG leads will be used to evaluate the width of QRS complex, where anyone with value of ≥120ms will be termed as having electrical dyssynchrony.

### Secondary outcomes

#### Worsening of heart failure

During the follow-up period, a patient with cardiac dyssynchrony and chronic heart failure on optimal medical tolerable therapy with no symptoms (stable) will be termed as having worsening of heart failure if signs and symptoms of decompensated heart failure develop and necessitates either escalation of oral diuretics dose or iv administration to relieve symptoms in the outpatient setting[25].

#### Readmission for Heart failure Symptoms

During follow-up, any worsening of heart failure symptoms in a patient with chronic heart failure and cardiac dyssynchrony that warrant in hospital management to relieve symptoms using diuretics and venodilators will be assessed as readmission for heart failure symptoms[25].

### Mortality

#### Cardiovascular death

Will be defined as any death that occurs following hospitalization due to heart failure or any sudden death with no any explainable cause in heart failure patients with cardiac dyssynchrony.

#### All-cause mortality

Any death occurring in a patient with heart failure and cardiac dyssynchrony hospitalized for any other disease condition apart from heart failure or death without an identifiable cause.

### Study variables

#### Aim 1 Study Variables

This variable addresses the prevalence of cardiac Dyssynchrony among patients with heart failure. The outcomes include electrical dyssynchrony that will be measured using an ECG assessing the width of the QRS complex. and mechanical dyssynchrony that will be assessed using echocardiography, both measured in milliseconds. Summary of description of aim 1, see table 1 below ….

**Table 1.** summary and description of objective number 1 of the study.

| variable | Method of measurement | Operational Definition of variable | Level of measurement |
| --- | --- | --- | --- |
| Cardiac dyssynchrony | Mechanical dyssynchrony will be assessed and measured using echo, while electrical dyssynchrony will be measured using 12 lead ECG | Cardiac dyssynchrony will be either mechanical, electrical or both<br>**Mechanical dyssynchrony** - will be termed as interventricular if the difference between LPEI and RPEI is 40ms. Intraventricular will be defined by the interval between the maximal outward displacement of the septum and the LV posterior wall delay being more than 130ms.<br>**Electrical dyssynchrony** - a standard 12 ECG leads will be used to evaluate the width of QRS complex, where anyone with a value of $\geq 120$ ms will be termed as having electrical dyssynchrony | Dichotomous |

#### Aim 2 Study Variables

These will assess the predictors of Cardiac dyssynchrony among heart failure patients. Age measured in years, ejection fraction measured in percentage, sex given as male or female, BP status given as high, low or normal, BMI given as obese or non obese, heart failure aetiology and symptoms of heart failure assessed using New York Heart Association Functional class. Summary of description of aim 2, see table 2 below.

**Table 2:** summary of description of predictors of outcome:

| Variable | Method of measurement | Operational definition | Level of measurements |
| --- | --- | --- | --- |
| Age | Counting the number of years from birth date (obtained from medical records) up to the date of enrollment. | Age in years | Continuous |
| Ejection fraction | Will be measured using ECHO. | Percentage of blood pumped out of the left ventricle after every beat | Continuous |
| Sex | Will be self-reported or inquired from medical records. | Male or female | Dichotomous |
| BMI | Will be calculated after measuring height using a stadiometer and weight using a calibrated weighing scale | Underweight<br><18.5kg/m<sup>2</sup><br>Normal 18.5-24.9kg/m<sup>2</sup><br>Overweight 25-29.9kg/m<sup>2</sup> | ordinal |
| | | Obese $\geq 30 \text{ kg/m}^2$ | |
| BP status | Medical records, self-reporting or assessment | High if $\geq 140/90 \text{ mmHg}$<br>Hypotensive if $< 90/60$<br>Normal if any value in between these readings | ordinal |
| Heart failure aetiology | ECHO | HHD: IF history of chronic hypertension or evidence of left ventricle hypertrophy on echo<br>IHD: if history of CAD, evidence of regional wall motion abnormalities on echo.<br>DCM: if evidence of left ventricular dilatation on echo<br>Co pulmonale: if there's coexistence of pulmonary diseases and right sided heart failure | nominal |
| Heart failure functional class | Symptoms: NYHA | **Class I (mild):** no symptoms with ordinary activities<br>**Class II (moderate):** | ordinal |
|  |  | symptoms on moderate<br>physical activity<br>**Class III (moderate to severe):** symptoms on<br>mild activities<br>**Class IV (severe):**<br>symptoms even at rest |  |

**Table 3:**
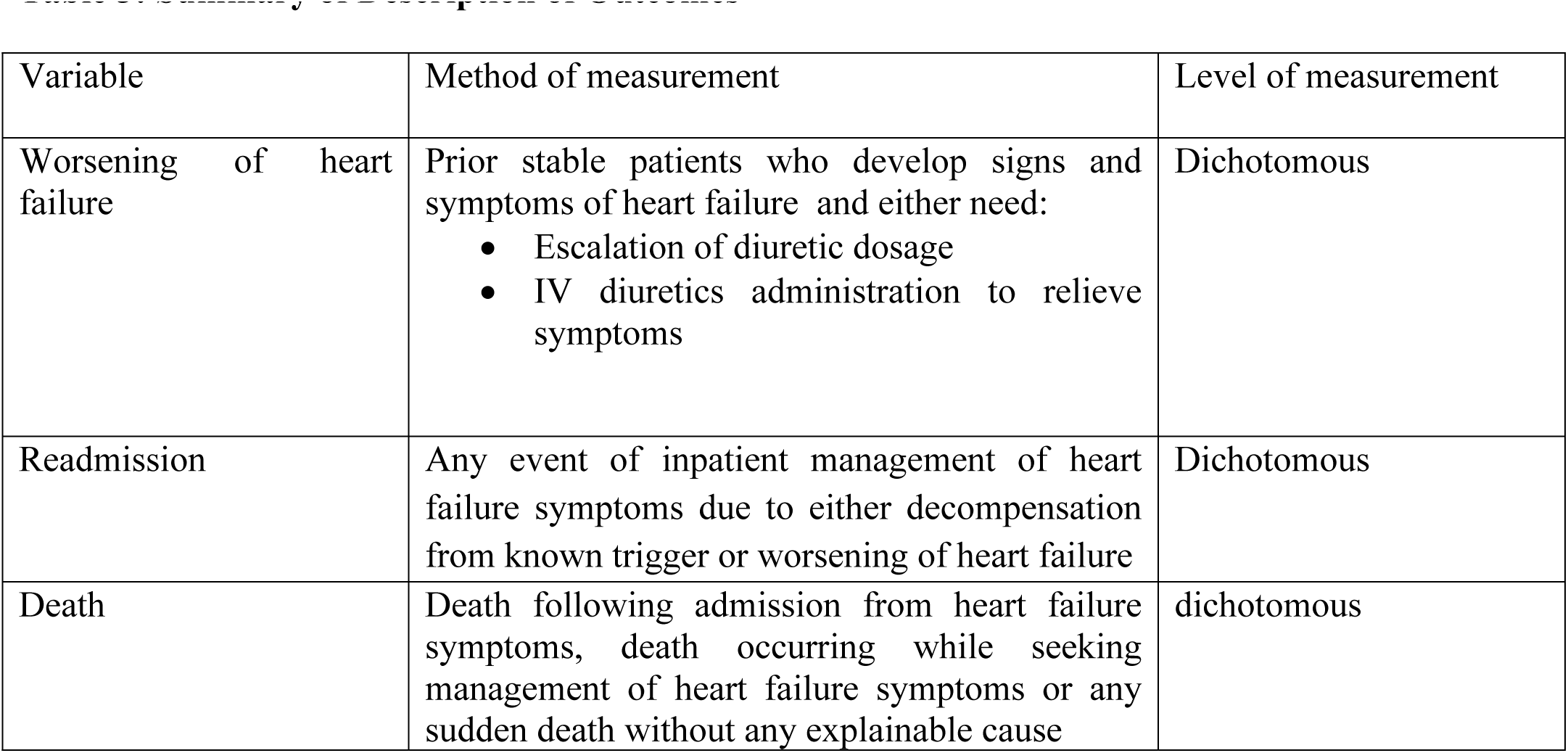
Summary of Description of Outcomes

#### Aim 3 study variables

these variables will assess the outcomes; readmissions, worsening of heart failure and death. Summary of description of aim 3, see table number 3 below.

### Participants’ characteristics

Participants will be adults aged 18 years or older who will be attending the cardiology department of the Benjamin Mkapa hospital with established diagnosis of heart failure using the framing harm criteria (two major criteria or one major and two minor) as described in the assessment of heart failure section.

### Data collection process

#### Evaluation of the participants

A total of 150 participants who will meet the inclusion criteria will be enrolled from July 2023. Participants who consent will be interviewed using a structured questionnaire that collects socio-demographic data such as age, gender, and area of residence. A minimum of two contacts; from the participant and next of kin, will be inquired. Moreover, alcohol consumption history will be taken with its duration (current drinker will be defined as alcohol consumption within the past 12 months, or currently taking alcohol while formerly used alcohol will be defined as someone who lastly took alcohol past 12months and currently doesn’t take alcohol)[1,26]. Cigarette smoking history will be taken and reported in cigarettes packs per year, the history of current smoking, will be defined as smoking within the past year or currently active smoking, while past smoker will be defined as someone who last smoke 12month ago and currently doesn’t smoke[1]. Information regarding potential predictors of cardiac dyssynchrony will also be collected, these include; heart failure symptoms functional class, presenting ejection fraction and conduction delay will be taken[27]. Also other clinical characteristics including history of Hypertension (defined as a history of Hypertension or the use of antihypertensive medications) and diabetes (defined as a history of diabetes or the use of diabetic medications) will be documented[28]

### Clinical variable measurement

#### Blood pressure measurement

A blood pressure (BP) reading will be measured using an automated digital machine of the AD Medical Inc. brand, keeping with the 2018 AHA/ACC Hypertension guideline for standard measurement of BP[29]. Hypertension will be defined as BP ≥140/90 mmHg after three different reading measured at least 5 minutes apart or a patient with a known history of hypertension or a patient on antihypertensive medications[30].

#### Radial pulse measurement

Two operators will assess the radial pulse using a timer for one minute. The arm of the participant will be slightly pronated and the wrist slightly flexed[31]. Radial pulses will be palpated against the radial head using the ring, middle and index fingers, in supinated[31]; To eliminate bias, the two operators will measure separately, and if they get separate values, an average of the two will be taken and us[32]

### Data management

Data will be collected using a structured questionnaire that was adopted and customized from a tool that was used in a multi-national heart failure registry; The global congestive heart failure (GCHF)[1]. Each participant will be assigned a unique anonymous identifier number. Data will be stored on password-protected and encrypted computers to maximize confidentiality and security. Anyone who will need to access the data for secondary data analysis an approval will be given by the authorized research board of the institution where the data will be stored in consultation with the primary researcher of this study.

### Data analysis

Data will be entered on a Microsoft Excel sheet for statistical analysis and then converted to IBM SPSS PC version 26. Continuous variables will be summarized as mean and standard deviation (SD), or median and interquartile ranges, for non-parametric variables; frequencies and percentages will be used for categorical variables. Descriptive statistics will be used to summarize demographics and clinical characteristics using frequencies and proportions. Binary logistic regression will be used to determine the predictors of outcome, and crude odds ratio will be used as a measure of association. The predictors for multivariable logistic regression will be those that are statistically significant in univariate logistic regression, those that literatures support that they have association with the outcome, or whose p value is ≤0.2.[33] These will include ejection fraction, male sex, NYHA, BMI, DM and BP status; and adjusted odds ratio will be used as a measure of association. At the same time, 95% confidence interval (CI) will be calculated concurrently and a two-sided p≤ 0.05 will be used to establish statistical significance.

### Ethical issues

The Vice Chancellor’s office at the University of Dodoma provided permission to conduct the study after obtaining ethical clearance from the Directorate of Research and Publications with reference number MA.84/261/02/’A’/59/14. Following that, the administrative department of Benjamin Mkapa approved the data collection with reference number AB.150/293/01/298. Participants will be informed that their participation is fully optional and that they may opt out at any moment without affecting their daily routine care. To ensure privacy and confidentiality, participants’ names are substituted with identification numbers; however, their standard of care is unaffected by their decision to participate or not. Those with cardiac dyssynchrony will be channelled to appropriate centres capable of resynchronization therapy and any other appropriate management that might benefit them.

#### Dissemination of results

The study’s findings are expected to be published in peer-reviewed journal the PLOS ONE with the aim of communicating these findings to the entire world to improve patient outcomes. Furthermore, these study findings will further be presented to the Ministry of Health to equip them with a basis for incorporating CRTDs as a must therapeutic service in our local guidelines to manage patients with heart failure and cardiac dyssynchrony.

### Study timeline

The study is expected to take twelve months; three months is expected to suffice participant enrollment, six months of follow-up and the remaining three months for data analysis and reporting of results.

## DISCUSSION

The presence of CD in HF patients has been linked to increased mortality rates and recurrent hospital admissions[34]. Studies have reported a 5-year overall survival rate of only 50% in HF patients[3] and a mortality rate of 68% in patients with HF and cardiac dyssynchrony after a 4-year follow-up[6]. Despite the introduction of novel pharmacologic medications like beta-blockers and angiotensin-converting-enzyme inhibitors (ACEIs) in HF management, the quality of life for these patients remains poor with the majority of them occurring as early as within 6-month.[34,35]. CD exacerbates the risk of death and worsens the survival outcomes in HF patients[9]. The study highlights the significant impact of cardiac dyssynchrony (CD) on patients with heart failure (HF) and its association with poor prognosis and reduced overall survival[34]. It emphasizes the need for timely diagnosis and appropriate treatment of CD to improve patient outcomes and quality of life. Interestingly, studies have shown that even though these people have severe heart failure, if they receive cardiac resynchronization therapy (CRT), they do better, have a lower risk of recurrent hospitalization, and lower mortality rate.

However, it can be challenging to stay in touch with every cohort member in a prospective cohort research, especially if the cohort is sizable and the time of follow-up is lengthy. When numerous patients are lost to follow-up, the results may be skewed, particularly when the risk factor or outcome of interest is connected to the reason for the loss to follow-up. Furthermore, the ability of a cohort study to deduce causality from the observed link between cardiac dyssynchrony and bad outcomes is limited.

The study site, Benjamin Mkapa Hospital, and the University of Dodoma library will get the final results, and a manuscript will be prepared for submission to a number of peer-reviewed journals prior to publication.

## Data Availability

Deidentified research data will be made publicly available when the study is completed and published.

## Acknowledgement

I would like to express my gratitude to my supervisors, Dr. John R. Meda, Dr. Azan Nyundo, and Dr. Baraka Alphonce for their tireless and endless guidance and support throughout the development of this study protocol. They had a significant role in forming the study topics and techniques with their work, knowledge, and insight. They have been steadfast in their dedication to assuring the accuracy and reliability of this study, and I appreciate how eager they have been to offer suggestions and support along the road. They provided the direction that made this protocol practicable.

## Author Contributions

**Conceptualization**: Patrick Bilikundi, John R. Meda

**Data curation**: Patrick Bilikundi

**Formal analysis**: Patrick Bilikundi

**Investigation**: John Meda

**Methodology**: Patrick Bilikundi, Baraka Alphonce Azan Nyundo, John Meda.

**Supervision**: Baraka Alphonce, Azan Nyundo, John Meda

**Writing - original draft**: Patrick Bilikundi

**Writing- review & editing**: Baraka Alphonce, Azan Nyundo, John Meda

